# Timing COVID-19 – Synchronization of longitudinal patient data to the underlying disease progression using CRP as a temporal marker

**DOI:** 10.1101/2020.06.11.20128041

**Authors:** Martina A. Maibach, Ahmed Allam, Matthias P. Hilty, Nicolas A. Perez Gonzalez, Philipp K. Buehler, Pedro D. Wendel Garcia, Silvio D. Brugger, Christoph C. Ganter, The CoViD-19 ICU-Research Group Zurich, The RISC-19-ICU Investigators, Michael Krauthammer, Reto A. Schuepbach, Jan Bartussek

## Abstract

Advances in medical technology and IT infrastructure have led to increased availability of continuous patient data that allows investigation of the longitudinal progression of novel and known diseases in unprecedented detail. However, to accurately describe any underlying pathophysiology with longitudinal data, the individual patient trajectories have to be synchronized based on temporal markers. In this study, we use longitudinal data from 28 critically ill ICU COVID-19 patients to compare the commonly used alignment markers “onset of symptoms”, “hospital admission” and “ICU admission” with a novel objective method based on the peak value of inflammatory marker C-reactive protein (CRP). By applying our CRP-based method to align the progression of neutrophils and lymphocytes, we were able to define a pathophysiological window that allowed further risk stratification in our COVID-19 patient cohort. Our data highlights that proper synchronization of patient data is crucial to differentiate severity subgroups and to allow reliable interpatient comparisons.

## Introduction

The rapid spread of corona virus disease 19 (COVID-19) imposes a heavy burden on public health systems around the world. A substantial number of patients show a severe disease progression possibly caused by endotheliitis, gas diffusion impairment and organ ischemia [1, 2]. Current research efforts focus on the identification of predictive indicators that allow closer supervision and targeted intervention in high-risk patients. As a hyper-activated immune response might act as a driving factor for severe COVID-19 progression [1], ratios between neutrophils and lymphocytes (NLR) [3], lymphocyte counts alone [4] and elevation of specific cytokines among other laboratory values [3, 5-7] have been proposed as markers for initial patient risk assessment and stratification. Currently, most studies solely compare measurements taken at hospital or intensive care unit (ICU) admission, neglecting the enormous potential of continuous longitudinal data obtained throughout hospitalization. This is especially detrimental for the most severe patients, as this high mortality risk group could benefit the most from a more detailed separation into different disease progression subgroups. However, the pooling of longitudinal data requires a temporal marker to align individual patient trajectories. In this pilot study, we compare different alignment methods and show that C-reactive protein (CRP) can be used to synchronize the individual patient trajectories with the underlying pathophysiology.

## Results

To date, only few studies show longitudinal data of COVID-19 patients, and single time point patient comparisons are often made based on clinical parameters such as onset of symptoms, hospital or ICU admission [3-7]. Alignment based on these values could potentially introduce unnecessary interpatient variation due to human bias and circumstantial parameters such as hospital capacity, resource availability or accessibility, and could eventually result in a clouding of the true underlying disease progression. Comparison of individual patient trajectories in our cohort of 28 critically ill COVID-19 patients admitted to the ICU of the University Hospital Zurich (Suppl. Table 1) already revealed considerable inter-patient variability (Fig. 1 a): Out of the 28 patients, 8 were directly transferred to the ICU upon hospital admission and 5 additional patients were transferred to the ICU only one day after hospital admission. Based on this data alone, it is evident that interpatient comparison at hospital or ICU admission was biased in our ICU COVID-19 patient cohort. Likewise, onset of symptoms showed a high variation (7.65 ± 8.49 days) and was occasionally missing. Group comparisons based on these clinical markers might result in the description of false differences, for example by comparing patients in late disease stages with patients in early disease stages, or occlusion of actual differences due to temporal misalignment (Fig. 1 b). These problems are encountered by most medical centers and researchers alike and highlight the necessity for an objective synchronization marker that aligns individual patient trajectories to the underlying pathophysiology.

**Fig. 1.**
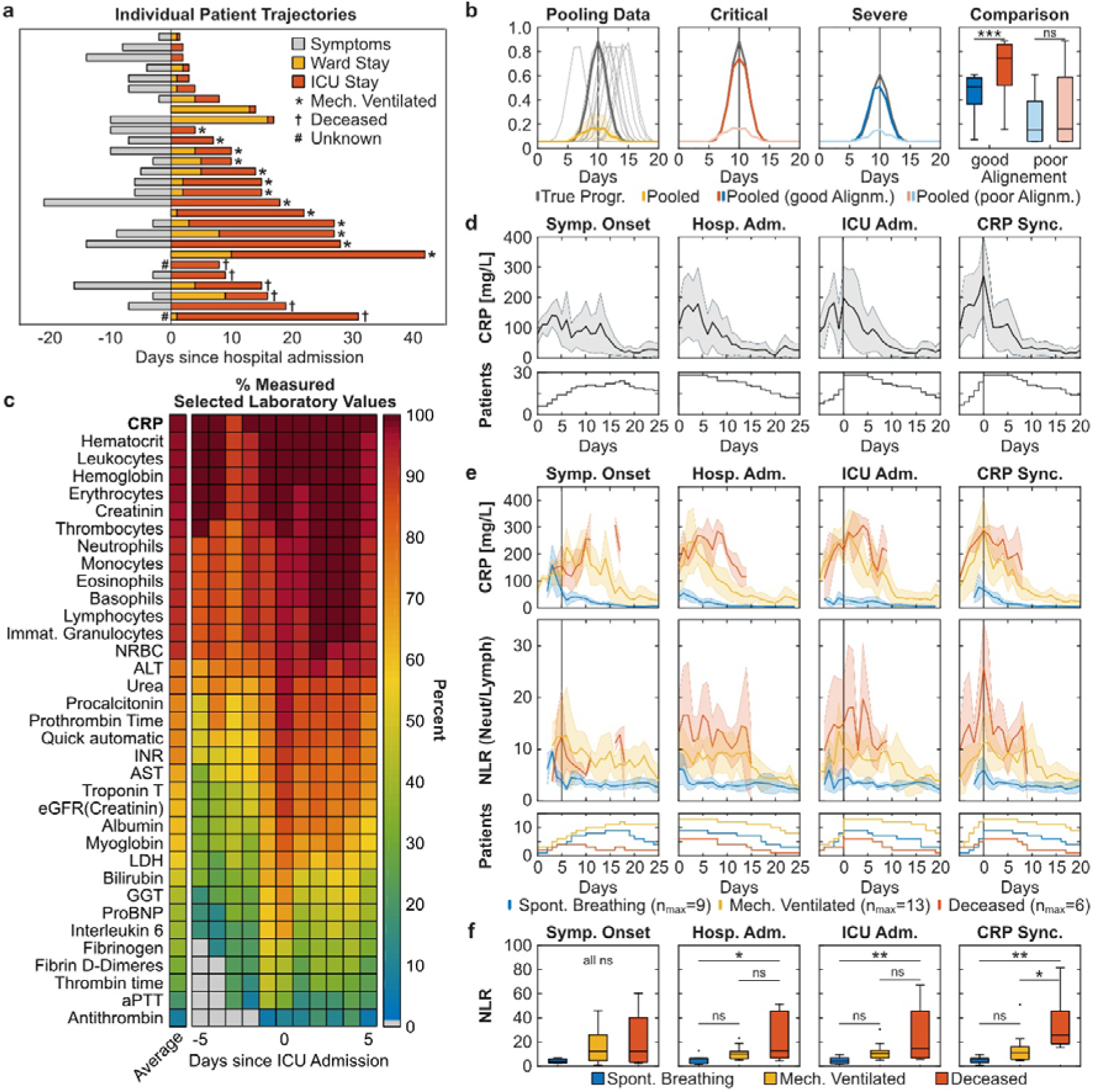
Pathophysiological synchronization of COVID-19 trajectories improves subgroup distinction. **a** Individual patient trajectories of 28 severe ICU COVID-19 cases. **b** Simulation illustrating the effect of pooling temporally shifted data. Left panel: Simulation of 100 identical exponential curves scattered around day 10 (normally distributed, σ=1.5, one sampling per day). Dark grey line: true progression without temporal scatter; light grey lines: 10 randomly selected curves of the simulation; yellow line: median ± MAD of the 100 simulated curves. Middle panels: Grey lines represent two identical curves that differ in height by 50%. Light colored curves: σ=1.5, dark colored curves σ=0.75. Right panel: Boxplot comparison of the simulated curves in the middle panels at time point 10 days. **c** Heat plot of average measurement frequency around ICU admission. **d - e** Time course of CRP overall (d) and CRP and NLR in severity subgroups (e). Synchronization based on onset of symptoms resulted in the exclusion of two deceased patients due to unclear data. Data is shown as median ± MAD. Curves are cut-off when data of fewer than three patients was available. The respective patient numbers are shown in the bottom panels. **f** Subgroup comparison of each alignment method at time point 0 for CRP_max_-based, hospital and ICU admission-based and time point +5 days for onset of symptoms based alignment (indicated by the grey line in subfigure e). Multiple comparison testing with tukey post-hoc test was performed on single time points. ns = not significant, * p ≤ 0.05, ** p ≤ 0.01, *** p ≤ 0.001.

An ideal disease timer should provide both an early indication of disease progression and should be measured routinely in most hospital settings. Previous patient cohort studies have correlated the serum levels of the inflammatory cytokine interleukin 6 (IL-6), myoglobin and cardiac troponin at hospital admission with COVID-19 severity [6]. However, these values were not measured on a daily basis around ICU admission both in our cohort (38.2%, 59.7%, 62.7% respectively) and in the international RISC-19-ICU registry cohort of critically ill COVID-19 patients (14.6%, 9.6%, 30.0% in Switzerland and 15.3%, 6.7%, 28.2% internationally), thereby making them poor candidates for longitudinal alignment (Fig. 1 c). Instead, the acute phase inflammatory marker CRP was measured routinely around ICU admission both in our cohort (98.2%) and in the RISC-19-ICU registry cohort (86.9% in Switzerland, 74.8% internationally). CRP is under direct transcriptional control of IL-6, but shows a wider peak, making it more likely to be recorded by daily measurements and when the patient is hospitalized [8]. In contrast to other frequently measured laboratory values such as hematological cell counts or creatinine, we found that most patients had a distinct CRP maximum around ICU admission in our cohort, indicating a correlation with COVID-19 severity and progression (Fig. 1 d). Some patients showed further CRP maxima during their ICU stay, probably resulting from coinfections or secondary damage [8]. We hypothesize that the first local CRP maximum (CRP_max_) is primarily related to COVID-19 progression and can therefore be used to synchronize individual patient trajectories to the underlying pathophysiology. In our patient cohort, alignment based on CRP_max_ decreased both interpatient variability in the longitudinal CRP curve (Fig. 1 d) and the variability of other laboratory values such as total leukocyte and relative neutrophil and lymphocyte counts to a similar extent than the clinically based ICU admission alignment (Suppl. Fig. 1).

Our primary goal to accurately align longitudinal patient data was to further differentiate between the most severe ICU COVID-19 patients. To test whether CRP_max_-based synchronization improves patient stratification in our ICU patient cohort, we retrospectively defined three severity subgroups: (1) deceased ICU patients (n=6), (2) discharged ICU patients that had been mechanically ventilated (n=13) and (3) discharged ICU patients that had been spontaneously breathing while in the ICU (n=9). CRP peak values were more than threefold higher in the mechanically ventilated patient subgroups (mean±SD, 346±147 mg/L) as compared to the spontaneously breathing subgroup (99±74 mg/L), but did not differ from the deceased subgroup (338±106 mg/L) (Fig 1 e). This lack of distinction is reflected in all alignment methods. In accordance to current literature, we further assessed the longitudinal progression of relative neutrophils and lymphocyte counts (Suppl. Fig. 2) and the ratio thereof (NLR, Fig. 1 e) in the three severity subgroups [3, 4]. While both admission-based and CRP_max_-based alignment improved subgroup separation, only CRP_max_-based synchronization revealed a distinct NLR turning point, occurring simultaneously with CRP_max_, thereby providing a window for maximal subgroup distinction. A linear mixed effect model [9] employing subgroup and time as fixed effects and per-patient random slopes as random effects confirmed a difference between the subgroups and the measured time points in a window of ±4 days around CRP_max_ (p<0.01, Suppl. Table 1), whereas the time wise difference was not detected in the data shifted by ICU admission (Suppl. Table 2). Similarly, when comparing the subgroups in single time points of each alignment method, only the CRP_max_-based synchronization resulted in a significant difference between the two most severe patient subgroups (Fig. 1 f). These findings strengthen our CRP_max_- based synchronization as a potential tool for patient alignment and subgroup stratification.

In a last step, we explored whether different patient synchronization methods might have an impact on future outcome prediction using machine learning techniques (Suppl. Fig. 3). We generated two feature vectors for each patient containing the mean values of CRP, relative neutrophils and lymphocytes of a time window anchored on either ICU admission or on CRP_max_ (Suppl. Fig. 3 upper panel). Using a stratified 5-fold cross validation logistic regression model, we found that the CRP_max_ anchoring increased the overall prediction accuracy by 9.6% and F1-macro score by 51.8% (accuracy 0.68 ± 0.22, F-score 0.668 ± 0.23) as compared to the ICU admission anchoring (accuracy 0.62 ± 0.10, F-score 0.44 ± 0.13) (Suppl. Fig. 3 b-c, window 1). Similarly, the corresponding confusion matrices indicated a higher accuracy in distinguishing between the most severe subgroups of mechanically ventilated and deceased ICU patients (Suppl. Fig. 3 d-e). Collectively, this data suggests that pathophysiological synchronization of longitudinal patient data has the potential to improve mortality-risk stratification and subgroup distinction of severe ICU patients both in a clinical setting and for research purposes.

## Discussion

In this pilot study containing 28 critically ill COVID-19 patients, we demonstrated that longitudinal data synchronization based on the inflammatory marker CRP reduces interpatient variability at least to an equal extend as the ICU admission based alignment. Both “onset of symptoms” and “hospital admission” were poor temporal markers, leading to increased variability, occlusion of subgroup differences and, in case of “onset of symptoms”, to patient exclusions due to unclear data. The interpretation and translation of noteworthy symptoms from patients to clinicians make “onset of symptoms” a highly subjective value for patient synchronization, which is reflected in our data and early reports of exaggerated incubation periods until onset of disease [5, 10]. While ICU admission is a consistent clinical marker in our monocentric study, this might not be the case when comparing patients from different hospitals with less stringent or deviating ICU admission criteria, resources or ICU capacity. Furthermore, COVID-19 associated symptoms might not be the primary reason for ICU admission in some patients and, obviously, this temporal marker cannot be applied to non-ICU patients. In contrast, our findings suggest that CRP can serve as an objective synchronization marker that allows alignment of disease trajectories independent of hospital specific policies, which is of special value for multicenter studies.

In line with previous studies, our subgroup analysis of both CRP_max_ and ICU aligned data reproduced the COVID-19 severity markers: neutrophilia, lymphocytopenia and the ratio thereof [3-7]. However, only CRP_max_-based longitudinal alignment improved distinction between the most severe subgroups of mechanically ventilated patients and deceased patients. Although this pilot study relies on a small cohort, our data suggests a central role for CRP in the timing of COVID-19 immunopathology by marking the turning point of longitudinal NLR dynamic and thereby providing a window for maximal subgroup distinction. Interestingly, CRP itself has immune-modulating functions such as complement activation, regulation of apoptosis and cellular processes of both neutrophils and monocyte-derived cells [8]. Although an elevation of CRP is generally associated with bacterial rather than viral infections [8, 11, 12], elevated CRP levels have been observed in COVID-19 patients as well as in severe progression of other respiratory viral diseases such as influenza [3, 4, 6, 13, 14]. It is tempting to speculate that elevation of CRP in severe respiratory viral infections marks a shift from a more localized inflammation of the lungs to a multi-organ systemic immune response.

Digitalization of modern medicine has led to increased availability of continuous patient data that should be used to describe and define longitudinal disease progression and pathophysiology of novel and known diseases alike. Our data highlights that proper synchronization of longitudinal patient trajectories to the underlying pathophysiology is crucial to differentiate severity subgroups and allow reliable interpatient comparisons.

## Data Availability

The data analyzed in anonymized form and code generated during the current study are available from the corresponding author on reasonable request. The RISC-19-ICU data set (NCT04357275) is available on reasonable request (https://www.risc-19-icu.net/main-page/how-to-participate).

## Acknowledgements

We thank Catharina Giese, Patrick Hirschi, the RDSC and the PDMS group of the University Hospital Zurich for their continued support. We further thank all the public health and essential workers as well as researchers for their efforts in the battle against SARS-CoV-2. This research was supported from non-restricted grants to Reto A. Schuepbach.

## Data Availability Statement

The datasets analyzed and code generated during the current study are available from the corresponding authors on reasonable request. The RISC-19-ICU data set (NCT04357275) is available on reasonable request (https://www.risc-19-icu.net/main-page/how-to-participate).

## Author Contributions

M.A.M. and J.B. conceptualized and performed the research and wrote the manuscript. A.A. performed the 5-fold cross-validation modelling. M.P.H performed the mixed linear regression modelling and RISC-19-ICU registry data analysis. The CoViD-19 ICU-Research Group Zurich and the RISC-19-ICU Investigators contributed to the data collection. A.A., S.D.B., P.K.B., C.C.G., N.A.P.G., M.P.H., M.K., R.A.S. and. P.D.W.G. provided valuable input and critically assessed the manuscript.

## The CoViD-19 ICU-Research Group Zurich

University Hospital of Zurich, Institute for Intensive Care: *Bartussek, Jan; Buehler, Phillip; Heuberger, Dorothea Monika; Hilty, Matthias Peter; Hofmänner, Daniel Andrea; Maibach, Martina Anna; Schuepbach Reto Andreas; Wendel Garcia, Pedro David*. University Hospital of Zurich, Department of Infectious Diseases and Hospital Epidemiology: *Brugger, Silvio; Mairpady Shambat, Srikanth; Zinkernagel, Annelies*

## The RISC-19-ICU Investigators

Separate list provided.

---

**Supplementary Figure 1.**
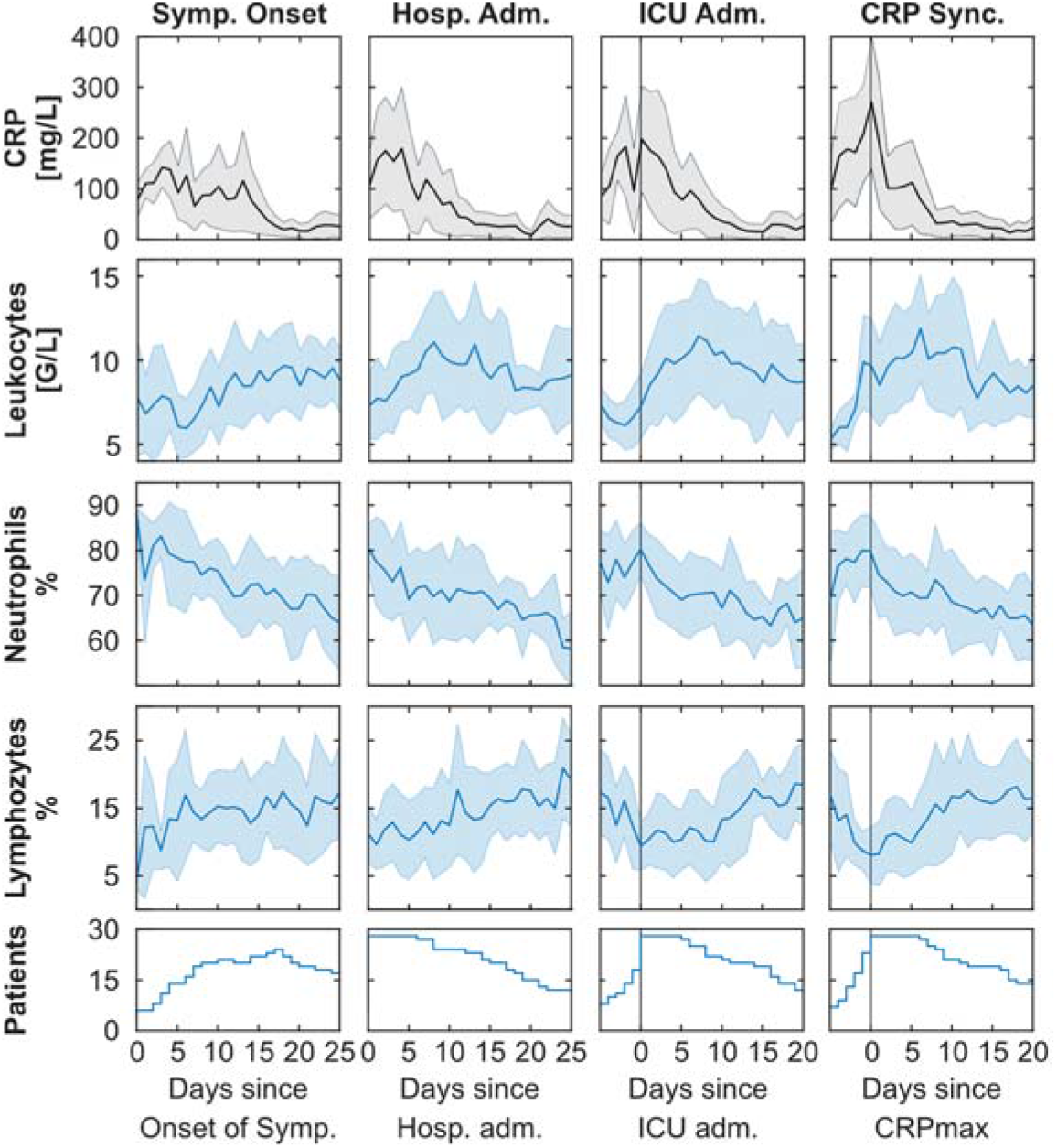
Dynamic changes of CRP, leukocytes and relative neutrophils and lymphocytes. Longitudinal CRP, leukocyte, neutrophils and lymphocytes shifted relative to symptom onset (left), hospital admission (middle, left), ICU admission (middle, right) or first local CRP maximum (right). Synchronization based on onset of symptoms resulted in the exclusion of two deceased patients due to unclear data. Data is shown as median ± MAD. Curves are cut-off when data of fewer than three patients was available. The respective patient numbers are shown in the bottom panels.

**Supplementary Figure 2.**
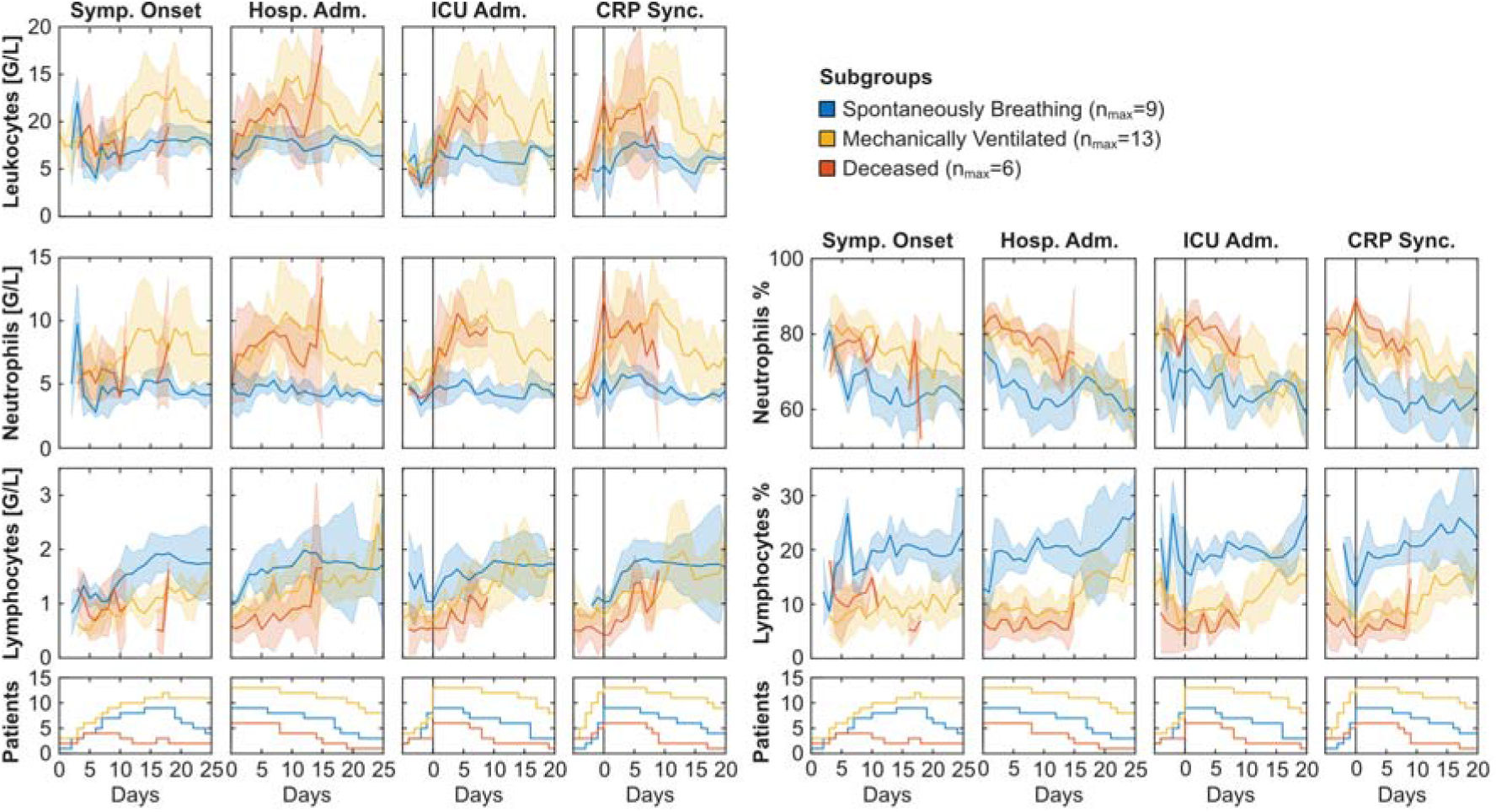
Dynamic changes of leukocytes, relative and absolute lymphocytes and neutrophils of severity subgroups. Longitudinal data is synchronized based on onset of symptoms (left) hospital admission (middle, left), ICU admission (middle, right) or first local CRP maximum (right). Synchronization based on onset of symptoms resulted in the exclusion of two deceased patients due to unclear data. Data is shown as median ± MAD. Curves are cut-off when data of fewer than three patients was available. The respective patient numbers are shown in the bottom panels.

**Supplementary Figure 3.**
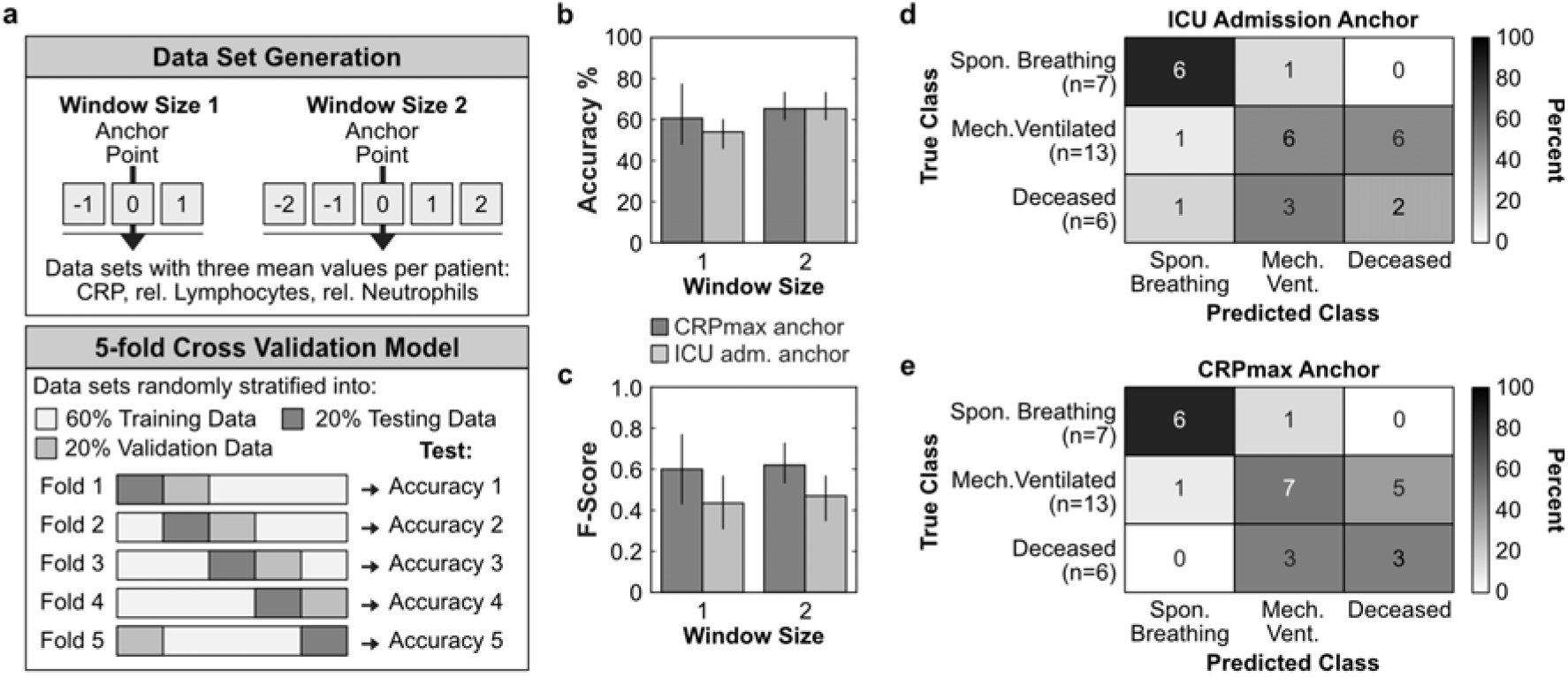
Timer-based risk stratification could improve outcome prediction. **a** Graphical representation of the data set generation and the applied 5-fold cross validation model. Two feature vectors were generated for each patient containing the mean values of CRP, relative neutrophils and lymphocytes of a time window anchored on either ICU admission or on CRP_max_ (upper panel). We followed a stratified 5-fold cross-validation scheme, where each fold was defined as a distinct 80%-20% train-test split. Within each fold, hyper-parameter selection was performed in the training set with a stratified 4-fold cross validation. For each fold multiple logistic regression models were trained using varying hyper-parameters such as regularization type (l1, l2) [15], regularization value in the interval of [10^−4^ – 10^4^], optimizer (LBFGS [16], SAGA [17]) and with or without class weighting. The best models as determined by F1-macro score on the 4-fold cross validation were then tested on the test split. Since our retrospective patient classification was done at ICU discharge, only values before outcome classification were considered for the construction of the feature vectors. This lead to the exclusion of 2 patients from the spontaneously breathing subgroup. **b – c** Mean accuracy performance (b) and mean Macro-f1 score (c) of ICU admission or CRP_max_ anchoring. Data is reported as mean ± SD. **d – e** Confusion matrices constructed from the best performing trained model of each fold using the test data from all five folds of the ICU admission anchored (d) or CRP_max_ anchored (e) window size 1 data set.

**Supplementary Table 1.** Patient characteristics at ICU admission. Non-binary data is shown as mean (SD) and binary data as mean (%). p-Values indicate one-way ANOVA for normally distributed data, Kruskal-Wallis test for non-normally distributed data and χ2-test for binary data.

|  | **Overall**<br>**(n=28)** | **Spont.**<br>**Breathing**<br>**(n=9)** | **Mech. Vent**<br>**(n=13)** | **Deceased**<br>**(n=6)** | **p** |
| --- | --- | --- | --- | --- | --- |
| **age,**<br>mean (SD) | 65.5 (11.4) | 67.8 (12.8) | 62.6 (9.5) | 68.5 (13.3) | 0.46 |
| **male sex,**<br>n (%) | 0.8 (0.4) | 0.7 (0.5) | 0.8 (0.4) | 1.0 (0.0) | 0.26 |
| **BMI,**<br>mean (SD) | 28.1 (4.2) | 30.3 (5.5) | 27.4 (3.4) | 26.4 (2.0) | 0.15 |
| **coronary artery disease,**<br>n (%) | 17 (60.7) | 4 (44.4) | 8 (61.5) | 5 (83.3) | 0.32 |
| **chronic heart failure,**<br>n (%) | 8 (28.6) | 2 (22.2) | 3 (23.1) | 3 (50.0) | 0.42 |
| **peripheral artery disease,**<br>n (%) | 1 (3.6) | 1 (11.1) | 0 (0.0) | 0 (0.0) | 0.33 |
| **arterial hypertension,**<br>n (%) | 9 (32.1) | 1 (11.1) | 5 (38.5) | 3 (50.0) | 0.23 |
| **diabetes mellitus,**<br>n (%) | 5 (17.9) | 2 (22.2) | 2 (15.4) | 1 (16.7) | 0.92 |
| **insulin dep. diabetes mellitus,**<br>n (%) | 11 (39.3) | 3 (33.3) | 4 (30.8) | 4 (66.7) | 0.30 |
| **symptoms to hosp. time,**<br>mean (SD) | 5.6 (5.0) | 5.9 (4.8) | 5.2 (5.8) | 6.2 (4.5) | 0.92 |
| **Hosp. to ICU adm. time,**<br>mean (SD) | 3.0 (5.0) | 3.9 (6.2) | 3.2 (5.2) | 1.3 (1.8) | 0.63 |
| **ICU length of stay,**<br>mean (SD) | 12.6 (11.3) | 8.4 (17.0) | 14.5 (7.9) | 13.8 (8.8) | 0.53 |
| **Apache II,**<br>mean (SD) | 17.5 (8.0) | 11.8 (7.0) | 18.8 (6.9) | 23.3 (6.6) | 0.01 |
| **SAPS II,**<br>mean (SD) | 57.5 (19.9) | 43.2 (18.1) | 60.6 (17.5) | 72.2 (15.4) | 0.01 |
| **SOFA,**<br>mean (SD) | 13.0 (4.6) | 9.9 (4.7) | 14.2 (4.1) | 15.0 (3.5) | 0.04 |

**Supplementary Table 2.** Mixed linear regression model for ICU shifted data. *P-Value indicate likelihood ratio test for overall analysis and Satterthwaite approximation for subgroup analysis*.

| NLR $\mu \pm$ SD, | Spont. Breathing (n=9) | Mech. Vent (n=13) | Deceased (n=6) | Time estimates ± S.E. (CI) | Degrees of freedom | p time |
| --- | --- | --- | --- | --- | --- | --- |
| day -4 | - | 9.87 ± 8.84 | 28.19 ± 32.41 | | | |
| day -2 | 6.34 ± 4.58 | 10.33 ± 5.88 | 30.06 ± 27.67 | 22.32 ± 6.04<br>(12.15 - 32.49) | 92.36 | 0.222 |
| day 0 | 4.79 ± 2.36 | 16.46 ± 19.83 | 38.22 ± 37.1 | 7.7 ± 3.3<br>(2.21 - 13.16) | 92.65 | **0.022** |
| day 2 | 3.87 ± 1.16 | 10.22 ± 6.99 | 23.54 ± 22.36 | 1.36 ± 3.3<br>(-4.12 - 6.83) | 92.65 | 0.681 |
| day 4 | 3.92 ± 1.67 | 10.23 ± 5.11 | 14.21 ± 13.3 | -0.62 ± 3.3<br>(-6.1 - 4.85) | 92.65 | 0.852 |
| Group estimates ± S.E. (CI) | | 7.2 ± 4.98<br>(-1.18 - 15.59) | 22.32 ± 6.04<br>(12.15 - 32.49) | | | **< 0.01** |
| Degrees of freedom |  | 29.48 | 29.19 |  |  |  |
| p group |  | 0.159 | **< 0.001** | **< 0.01** |  |  |

**Supplementary Table 3.** Mixed linear regression model for CRP_max_ shifted data. P-Value indicate likelihood ratio test for overall analysis and Satterthwaite approximation for subgroup analysis.

| NLR $\mu \pm$ SD, | Spont. Breathing (n=9) | Mech. Vent (n=13) | Deceased (n=6) | Time estimates ± S.E. (CI) | Degrees of freedom | p time |
| --- | --- | --- | --- | --- | --- | --- |
| day -4 | 5.99 ± 4.1 | 8.13 ± 4.72 | 33.01 ± 42.28 | | | |
| day -2 | 5.26 ± 3.79 | 12.77 ± 5.91 | 30.16 ± 36.4 | -0.97 ± 3.65<br>(-7.01 - 5.08) | 85.10 | 0.791 |
| day 0 | 4.41 ± 1.97 | 14.36 ± 14.66 | 30.69 ± 33.53 | 1.19 ± 3.48<br>(-4.59 - 6.96) | 86.85 | 0.734 |
| day 2 | 3.57 ± 0.96 | 13.19 ± 12.52 | 17.08 ± 12.81 | -2.55 ± 3.48<br>(-8.33 - 3.22) | 86.85 | 0.466 |
| day 4 | 4.01 ± 1.65 | 11.62 ± 11.83 | 24.03 ± 18.56 | -1.75 ± 3.52<br>(-7.58 - 4.08) | 87.17 | 0.620 |
| Group estimates ± S.E. (CI) | | 7.87 ± 4.96<br>(-0.5 - 16.21) | 20.93 ± 6.02<br>(10.77 - 31.07) | | | 0.600 |
| Degrees of freedom |  | 28.46 | 28.44 |  |  |  |
| p group |  | 0.124 | **0.002** | **< 0.01** |  |  |

**RISC-19-ICU Investigators**

**Andorra:** Unidad de Cuidados Intensivos, Hospital Nostra Senyora de Meritxell, Escaldes-Engordany (Mario Alfaro Farias, MD; Antoni Margarit, MD; Gerardo Vizmanos-Lamotte, MD). **Austria:** Department for Anesthesiology and Intensive Care, Johannes Kepler University Linz, Linz (Thomas Tschoellitsch, MD; Jens Meier, MD); Dept. Of Pediatrics, Medical University Vienna, Vienna (Francesco S. Cardona, MD, MSc). **Czech Republic:** Klinika anesteziologie perioperacni a intenzivni mediciny, Masaryk Hospital, Usti nad Labem (Josef Skola, MD; Lenka Horakova, MD). **Ecuador:** Unidad de Cuidados Intensivos, Hospital Vicente Corral Moscoso, Cuenca (Hernan Aguirre-Bermeo, MD, PhD; Janina Apolo, BSc). **France:** Department of Anesthesiology and Critical Care Medicine, University Hospital of Nancy (Emmanuel Novy, MD; Marie-Reine Losser, MD, PhD). SCPARE- Intensive Care Unit, Clinique Louis Pasteur, Essey-lès-Nancy (Geoffrey Jurkolow, MD; Gauthier Delahaye, MD). **Germany:** Medical School Hannover, Medical Intensive Care, Hannover (Sascha David, MD; Tobias Welte, MD); Department of Medicine III - Interdisciplinary Medical Intensive Care, Medical Center University of Freiburg, Freiburg (Tobias Wengenmayer, MD; Dawid L. Staudacher, MD). **Greece:** Intensive Care Unit, St. Paul General Hospital of Thessaloniki, Thessaloniki (Theodoros Aslanidis, MD). **Hungary:** Departement of Anaethesiology and Intensive Care, University of Szeged, Hungary (Barna Babik, MD, PhD; Anita Korsos, MD); Department of Anaesthesia and Intensive Care, Semmelweis University, Budapest (Janos Gal, MD, PhD; Hermann Csaba, MD, PhD). **Italy:** Anesthesia and Intensive Care Unit, Azienda Ospedaliero Universitaria Ospedali Riuniti, Ancona (Abele Donati, MD, PhD; Andrea Carsetti, MD); Internal Medicine, Azienda Ospedaliera Universitaria di Modena, Modena (Fabrizio Turrini, MD, MSc; Maria Sole Simonini, MD); UO Anestesia e Terapia Intensiva, IRCCS Centro Cardiologico Monzino, Monzino (Roberto Ceriani, MD; Martina Murrone, MD); Department of Anesthesia and Intensive Care Medicine, Policlinico San Marco, Zingonia (Emanuele Rezoagli, MD, PhD; Giovanni Vitale, MD); Anesthesia and Intensive care, Azienda Ospedaliero-Universitaria di Ferrara, Cona (Alberto Fogagnolo, MD; Savino Spadaro, MD, PhD); Department of Internal Medicine, ASST Fatebenefratelli Sacco - “Luigi Sacco” Hospital, Milan (Maddalena Alessandra Wu, MD; Chiara Cogliati, MD); Division of Anesthesia and Intensive Care, ASST Fatebenefratelli Sacco - “Luigi Sacco” Hospital, Milan (Riccardo Colombo, MD; Emanuele Catena, MD); UOC Anestesia e Rianimazione, Ospedale Infermi, Rimini (Francesca Facondini, MD; Antonella Potalivo, MD); UO Pronto Soccorso Medicina d’Urgenza, Ospedale Infermi, Rimini (Gianfilippo Gangitano, MD; Tiziana Perin, MD); Department of Anesthesiology and Intensive Care Medicine, Fondazione Policlinico Universitario Agostino Gemelli IRCCS, Rome (Maria Grazia Bocci, MD; Massimo Antonelli, MD). **Netherlands:** Department of Intensive Care Medicine, Erasmus Medical Center, Rotterdam (Diederik Gommers, MD, PhD; Can Ince, PhD). **RISC-19-ICU board**: Matthias Peter Hilty, MD; Pedro David Wendel Garcia, MSc; Reto Andreas Schüpbach, MD; Jonathan Montomoli, MD, PhD; Philippe Guerci, MD; Thierry Fumeaux, MD. **Spain:** Servei de Medicina intensiva, Hospital Verge de la Cinta, Tortosa (Eric Mayor-Vázquez, MD); Servicio de Medicina Intensiva, Hospital Universitario de Torrejon, Madrid (Maria Cruz Martin Delgado, MD); Intensive Care, Complejo Hospitalario Universitario A Coruña, A Coruña (Raquel Rodriguez Garcia, MD; Jorge Gamez Zapata, MD); Unidad de Cuidados Intensivos, Hospital Clinico Universitario Lozano Blesa, Zaragoza (Begoña Zalba-Etayo, MD, PhD; Herminia Lozano-Gomez, MD); Medical Intensive Care Unit, Hospital Clinic de Barcelona, Barcelona (Pedro Castro, MD; Adrian Tellez, MD); Anesthesiology Intensive Care Unit, Hospital Clinic de Barcelona, Barcelona (Adriana Jacas, MD; Guido Muñoz, MD); Acute Critical Cardiac Care Unit, Hospital Clinic de Barcelona, Barcelona (Rut Andrea, MD; Jose Ortiz, MD); Cardiovascular Surgery Critical Care Unit, Hospital Clinic de Barcelona, Barcelona (Eduard Quintana, MD; Irene Rovira, MD); Liver Intensive Care Unit, Hospital Clinic de Barcelona, Barcelona (Enric Reverter, MD; Javier Fernandez, MD); Respiratory Intensive Care Unit, Hospital Clinic de Barcelona, Barcelona (Miquel Ferrer, MD; Joan R. Badia, MD); Servicio de Medicina Intensiva, Hospital General San Jorge, Huesca (Arantxa Lander Azcona, MD; Jesus Escos Orta, MD). **Switzerland:** Institute of Intensive Care Medicine, University Hospital Zurich, Zurich (Philipp Bühler, MD; Silvio Brugger, MD, PhD; Daniel Hofmaenner, MD; Simone Unseld, MD; Frank Ruschitzka, MD; Jan Bartussek, Martina Maibach, PhD; PhD; Annelies Zinkernagel, MD, PhD); Interdisziplinaere Intensivstation, Spital Buelach, Buelach (Bernd Yuen, MD; Thomas Hillermann, MD); Soins Intensifs, Hopital cantonal de Fribourg, Fribourg (Hatem Ksouri, MD, PhD; Govind Oliver Sridharan, MD); Departement for intensive care medicine, Kantonsspital Nidwalden, Stans (Anette Ristic, MD; Michael Sepulcri, MD); Departement of Anesthesiology and Intensive Care Medicine, Cantonal Hospital St. Gallen, St. Gallen (Miodrag Filipovic, MD; Urs Pietsch, MD); Intensivstation, Regionalspital Emmental AG, Burgdorf (Petra Salomon, MD; Iris Drvaric, MD); Institut fuer Anesthaesie und Intensivmedizin, Zuger Kantonsspital AG, Baar (Peter Schott, MD; Severin Urech, MD); Intensivmedizin, St. Claraspital, Basel (Adriana Lambert, MD; Lukas Merki, MD); Department Intensive Care Medicine, Spitalzentrum Biel, Biel (Marcus Laube, MD); Intensivmedizin, Kantonsspital Graubünden, Chur (Frank Hillgaertner, MD; Marianne Sieber); Institut fuer Anaesthesie und Intensivmedizin, Spital Thurgau, Frauenfeld (Alexander Dullenkopf, MD; Lina Petersen, MD); Division of Neonatal and Pediatric Intensive Care, Geneva University Hospitals, Geneva (Serge Grazioli, MD; Peter C. Rimensberger, MD); Soins Intensifs, Hirslanden Clinique Cecil, Lausanne (Isabelle Fleisch, MD; Jerome Lavanchy, MD); Interdisziplinaere Intensivstation, Spital Maennedorf AG, Maennedorf (Katharina Marquardt, MD; Karim Shaikh, MD); Intensivmedizin, Schweizer Paraplegikerzentrum Nottwil, Nottwil (Hermann Redecker, MD); Intensivmedizin, Spital Oberengadin, Samedan (Michael Stephan, MD; Jan Brem, MD); Paediatric Intensive Care Unit, Children’s Hospital of Eastern Switzerland, St. Gallen (Bjarte Rogdo, MD; Andre Birkenmaier, MD); Klinik für Anaesthesie und Intensivmedizin, Spitalzentrum Oberwallis, Visp (Friederike Meyer zu Bentrup, MD, MBA); Interdisziplinaere Intensivstation, Stadtspital Triemli, Zurich (Patricia Fodor, MD; Pascal Locher, MD); Department Intensivmedizin, Universitaetsspital Basel, Basel (Martin Siegemund, MD; Nuria Zellweger); Department of Intensive Care Medicine, University Hospital Bern - Inselspital, Bern (Marie-Madlen Jeitziner, RN, PhD; Beatrice Jenni-Moser, RN, MSc); Intensivstation, Spital Grabs, Grabs (Christian Bürkle, MD); Medical ICU, Cantonal Hospital St.Gallen, St. Gallen (Gian-Reto Kleger, MD); Service d’Anesthesiologie, EHNV, Yverdon-les-Bains (Marilene Franchitti Laurent, MD; Jean-Christophe Laurent, MD); Abteilung für Anaesthesiologie und Intensivmedizin, Hirslanden Klinik Im Park, Zürich (Tomislav Gaspert, MD; Marija Jovic, MD); Intensivmedizin & Intermediate Care, Kantonsspital Olten, Olten (Michael Studhalter, MD); Institut für Anaesthesiologie und Intensivmedizin, Klinik Hirslanden, Zurich (Christoph Haberthuer, MD; Roger F. Lussman, MD); Anaesthesie Intensivmedizin Schmerzmedizin, Spital Schwyz, Schwyz (Daniela Selz, MD; Didier Naon, MD); Dipartimento Area Critica, Clinica Luganese Moncucco, Lugano (Romano Mauri, MD; Samuele Ceruti, MD); Institut für Anaesthesiologie Intensivmedizin & Rettungsmedizin, See-Spital Horgen & Kilchberg, Horgen (Julien Marrel, MD; Mirko Brenni, MD); Klinik für Operative Intensivmedizin, Kantonsspital Aarau, Aarau (Rolf Ensner, MD); Intensivstation, Kantonsspital Schaffhausen, Schaffhausen (Nadine Gehring, MD); Intensivstation, Spital Simmental-Thun- Saanenland AG, Thun (Antje Heise, MD); Klinik für Anaesthesie Intensivmedizin Operationszentrum und Schmerzmedizin, Kantonsspital Muensterlingen, Muensterlingen (Tobias Huebner, MD; Thomas A. Neff, MD); Division of Intensive Care, University Hospitals of Geneva, Geneva (Sara Cereghetti, MD; Filippo Boroli, MD; Jerome Pugin, MD, PhD). **United Kingdom:** Harefield Hospital, Royal Brompton & Harefield NHS Foundation Trust, Harefield (Nandor Marczin, MD, PhD; Joyce Wong, MD).

